# A Pharmacogenomic-Informed Representation Improves Multimodal EHR Survival Prediction

**DOI:** 10.64898/2026.01.27.26344981

**Authors:** Mun Hwan Lee, Yang Xiao, Xiaodi Li, Eric W. Klee, Ping Yang, Terence Sio, Liewei Wang, James Cerhan, Nansu Zong

**Affiliations:** Mayo Clinic, Rochester, Minnesota, USA

## Abstract

**Background:** Electronic health record (EHR)–based prognostic modeling is increasingly used in oncology, yet incorporating pharmacogenomic (PGx) knowledge derived from experimental systems into clinical prediction frameworks remains challenging. This gap is driven by fundamental mismatches between controlled drug–mutation assays and heterogeneous, incomplete real-world clinical data.

**Methods:** We propose a representation transfer framework that integrates PGx embeddings learned from large-scale in vitro pharmacogenomic screens into patient-level EHR models. A frozen pharmacogenomic encoder is used to generate interaction-aware embeddings from patient mutation profiles and administered therapies, which are aggregated into a fixed-length PGx Complementarity Representation. These representations are incorporated into multimodal survival prediction models alongside standard clinical features. Performance was evaluated using systematic modality ablation analyses, attribution analyses, and exploratory unsupervised representation analyses.

**Results:** Integrating PGx embeddings yielded consistent performance improvements across all evaluated modality combinations. Relative gains were largest in modality-sparse settings, where baseline EHR features encode limited biological context, and were attenuated—but remained significant—in biologically enriched configurations. Attribution analyses indicated that PGx embeddings contributed non-redundant predictive signal beyond standard clinical features. Exploratory unsupervised analyses further demonstrated that the learned representations exhibit interpretable association patterns aligned with known therapeutic exposures and pathway-level associations.

**Conclusion:** These findings suggest that externally learned pharmacogenomic representations can be transferred into real-world EHR models as a context-dependent, non-redundant augmentation. By framing PGx knowledge as an interaction-aware representation rather than a mechanistic model, this work provides an informatics framework for integrating experimental pharmacogenomic data into clinical prediction tasks in a reproducible and interpretable manner.

## 1. Introduction

Lung cancer remains the leading cause of cancer-related mortality worldwide (1), with prognosis varying widely across patients despite advances in molecular profiling and treatment selection (2). Accurate risk stratification is critical for guiding therapeutic decisions, prioritizing follow-up intensity, and identifying patients who may benefit from alternative treatment strategies (3, 4). In real-world clinical practice, however, prognosis is shaped by a complex interplay of biological, physiological, and therapeutic factors that are only partially captured in electronic health records (EHRs) (5, 6). As a result, survival prediction from EHR data remains a challenging task, even with the growing adoption of machine learning based approaches (7-10), which are ultimately constrained by the structure and granularity of clinically recorded variables.

Crucially, the clinical relevance of survival prediction relies heavily on the specific disease context. While long-term survival is the primary endpoint for early-stage disease, the clinical reality for real-world EHR cohorts is often dominated by advanced cases (11). In Stage IV non-small cell lung cancer (NSCLC) and extensive-stage small-cell lung cancer (SCLC), the median survival historically approximates one year (12, 13). This creates a critical “one-year horizon” that serves as a practical watershed for decision-making regarding treatment escalation, clinical trial eligibility, and palliative care planning (14). Therefore, in such high-risk settings, we define one-year mortality not as a simplified proxy, but as the most clinically grounded target for risk stratification.

Despite these high stakes, existing multimodal EHR models remain fundamentally constrained by the information explicitly recorded in clinical systems. EHRs document administered drugs and observed mutations as isolated variables. However, they fail to encode the mechanistic logic connecting these elements. Specifically, the data does not capture how genomic alterations dictate tumor sensitivity or resistance to therapy. (15). This omission creates a fragmented view of the patient, where the critical drug-gene interactions driving survival in advanced disease remain hidden within the data gaps (16). External pharmacogenomic resources, such as the Genomics of Drug Sensitivity in Cancer (GDSC), systematically characterize these interactions (17). Yet most existing survival prediction models have limited capacity to incorporate this external knowledge at the patient level, relying instead on surface level representations of mutations and treatments recorded in clinical systems (18).

This disconnect constrains the ability of EHR based models to capture the biological logic that links molecular alterations to therapeutic outcomes.

In this study, we address this gap by introducing a pharmacogenomics-informed embedding (PGx embedding), conceptualized as a PGx Complementarity Representation. This representation encodes the interaction-informed logic by which genomic alterations dictate therapeutic response. Unlike conventional embeddings that summarize observed clinical correlations, the PGx Complementarity Representation is explicitly trained to capture interaction-level pharmacogenomic mechanisms, enabling principled transfer across patients and treatment contexts. By learning latent drug–mutation interaction patterns from large-scale pharmacogenomic sensitivity data, we construct a compact representation that synthesizes fragmented clinical signals into a unified biological context.

We evaluated this framework in a real-world, molecularly profiled lung cancer cohort derived from tertiary care practice. Although modest in size (N=184), the cohort is heavily enriched for advanced metastatic disease (72.7%), representing the population for whom pharmacogenomic stratification is most clinically urgent. By prioritizing comprehensive gene panel testing, we ensured a high density of drug–mutation interactions and mortality events. This event-rich clinical context enables rigorous evaluation of prognostic modeling under clinically realistic constraints across diverse baseline architectures.

Our results demonstrate that the PGx embedding fundamentally reshapes model decision-making, emerging as the dominant predictive driver across architectures. The PGx Complementarity Representation delivers its largest relative benefit in data-sparse settings, effectively filling mechanistic gaps where biological context is otherwise missing, while exhibiting diminishing returns in biologically rich configurations. Furthermore, unsupervised analyses reveal that PGx embeddings spontaneously organize patients into biologically coherent phenotypes linking molecular pathways and therapeutic exposures. Together, these findings establish a scalable framework for precision oncology, demonstrating that frozen pharmacogenomic representations can reconstruct missing biological structure and improve outcome prediction in high-risk lung cancer.

## 2. Method

### 2.1 Overview of Study Design

We developed a two-stage framework to integrate external pharmacogenomic knowledge into multimodal EHR-based survival prediction, operationalizing the concept of a PGx Complementarity Representation within a controlled methodological setting. The central premise of this design is that mechanistic drug–mutation interaction information, which is not explicitly encoded in structured EHR data, can be learned from large-scale experimental pharmacogenomic resources and subsequently transferred to clinical prediction tasks.

In Stage 1 (Representation Learning), we trained a pharmacogenomic encoder using the Genomics of Drug Sensitivity in Cancer (GDSC) dataset (17), which systematically profiles drug response across diverse cancer cell lines. Using a large language model backbone, the encoder was trained to predict drug sensitivity from textualized representations of paired mutation profiles and drug structures. This stage was conducted exclusively on experimental cell-line data and did not incorporate any patient-level clinical or outcome information.

In Stage 2 (Representation Transfer), the learned pharmacogenomic representations were applied to a real-world lung cancer cohort. For each patient, a pharmacogenomic embedding was constructed based on observed somatic mutations and documented treatment exposures. Importantly, the encoder parameters were frozen during this stage, and no fine-tuning was performed using clinical outcomes. This design maintains a clear separation between representation learning and outcome prediction and allows the pharmacogenomic embedding to function as an externally derived feature.

Finally, patient-level PGx embeddings were concatenated with standard multimodal EHR features, including laboratory measurements, genomic indicators, and medication records, and used as inputs for downstream survival prediction models. This framework enables systematic evaluation of the contribution of externally learned pharmacogenomic information under controlled experimental conditions, while minimizing the risk of information leakage.

### 2.2 Pharmacogenomic Representation Learning (Stage 1)

To construct a pharmacogenomics-informed representation that captures latent drug–mutation interaction mechanisms independently of clinical outcomes, we trained a dedicated pharmacogenomic encoder using large-scale experimental data. This stage represents a key component of our framework, designed to extract latent mechanistic signals underlying drug-mutation interactions that are not explicitly encoded in routine EHR features.

#### 2.2.1 Pharmacogenomic Source Data

We leveraged the Genomics of Drug Sensitivity in Cancer (GDSC) dataset, which systematically profiles drug response across diverse cancer cell lines. Each training instance consisted of a drug–mutation pair associated with a binary sensitivity label (sensitive vs. resistant) obtained from the GDSC. Binary labels were defined by the original GDSC authors using drug-specific binarization thresholds optimized per compound based on pan-cancer IC50 distributions.

Drug information included both the standardized drug name and its SMILES string to encode chemical structure, while genomic context was represented by the set of somatic mutation gene symbols observed in the corresponding cell line. To focus on cancer-relevant genomic alterations, we restricted the mutation set to Hallmark cancer-related genes derived from the Molecular Signatures Database (MSigDB) hallmark gene set collection (19). This dataset is entirely experimental and devoid of patient-level clinical or survival information, ensuring strict separation from downstream clinical prediction tasks. Binary sensitivity labels were used to emphasize relative drug–mutation interaction patterns rather than absolute response magnitudes, enabling consistent learning across heterogeneous experimental conditions.

#### 2.2.2 Textual Encoding of Drug–Mutation Context

To enable joint modeling of heterogeneous pharmacogenomic inputs, we transformed structured drug and mutation data into unified textual prompts. This textualization allows the language model to encode relational associations between chemical structures and genomic alterations within a shared representation space. We employed the following structured prompt template:

> Instruction: Predict the sensitivity of the cancer cell line to the drug based on the following genomic and chemical context. Input: Drug: [Drug Name]; SMILES: [SMILES String]; Mutations: [Gene 1], [Gene 2], … Response:

#### 2.2.3 LLM-based Encoder Training via LoRA

We adopted TxGemma-2B (20), a publicly available large language model previously described for therapeutic-domain applications, as the backbone of our pharmacogenomic encoder. To efficiently adapt the model while preserving its pre-trained knowledge, we employed Low-Rank Adaptation (LoRA) (21). LoRA adapters were injected into the attention and feed-forward projection layers, while the majority of pre-trained parameters remained frozen. A linear classification head was attached to the final hidden state of the end-of-sequence (EOS) token, and the model was fine-tuned to minimize binary cross-entropy loss on the GDSC dataset.

#### 2.2.4 Finetuning Duration and Checkpoint Selection

To determine an appropriate finetuning duration, we evaluated multiple model checkpoints obtained at different finetuning steps. Model selection was guided not only by pharmacogenomic evaluation metrics observed during training, but also by downstream performance when the learned representations were transferred to patient-level cohort prediction tasks. This analysis informed the selection of a checkpoint that balanced pharmacogenomic task performance and downstream generalization. Based on this criterion, the checkpoint at 210,000 finetuning steps was selected for all subsequent analyses.

#### 2.2.5 PGx Embedding Extraction

Following Stage 1 training, the classification head was discarded, and the fine-tuned TxGemma-2B model was used as a feature extractor. For each drug–mutation input, we extracted the hidden representation of the final token from the last transformer layer, yielding a pharmacogenomic embedding vector of dimensionality 2,048 that captures latent interaction patterns learned during sensitivity prediction.

#### 2.2.6 Methodological Safeguards

After Stage 1, all encoder parameters were frozen, and no further fine-tuning was performed using clinical data or survival outcomes. This frozen representation transfer ensures that the PGx embedding functions as an external complementary signal when integrated into downstream EHR models, thereby minimizing the risk of outcome leakage or implicit proxy learning.

### 2.3 Patient-Level Embedding Construction (Stage2)

A central methodological challenge in transferring pharmacogenomic representations learned from experimental cell-line data to clinical applications lies in the mismatch of analytical units between the two domains. While the encoder trained in Stage 1 operates on discrete drug–mutation pairs, clinical patients are characterized by composite somatic mutation profiles and heterogeneous treatment histories involving multiple pharmacologic agents. This section describes the inference and aggregation strategy used to bridge this structural gap by constructing a single, fixed-length pharmacogenomic representation at the patient level.

#### 2.3.1 Zero-Shot Inference on Clinical Treatment

For each patient, we extracted the observed somatic mutation profile and all administered pharmacologic agents documented at or before the predefined landmark time. From these records, we generated Cartesian pairs consisting of the patient’s mutation profile and each administered drug possessing a valid chemical structure definition. Each drug–mutation pair was independently processed by the frozen pharmacogenomic encoder to generate a drug-specific embedding.

Importantly, patient-level PGx embeddings were generated without access to any clinical outcomes, follow-up duration, or post-landmark information. All inputs to the encoder were strictly limited to genomic alterations and medication exposures available at the landmark time, ensuring that embedding construction remained fully independent of survival outcomes and preventing implicit outcome leakage.

A key feature of this framework is the reliance on SMILES strings to represent chemical identity within the textualized input. Unlike vocabulary-based approaches restricted to a fixed set of training drugs, the language model processes intrinsic chemical syntax encoded in SMILES representations. This design allows the encoder to generate embeddings for clinical medications not explicitly observed during GDSC training by leveraging learned representations of chemical substructures in conjunction with genomic context. As a result, this approach supports representation-level generalization to a broader range of clinical drugs without requiring additional fine-tuning or outcome supervision.

#### 2.3.2 Aggregation into a Cumulative Pharmacogenomic Context

The inference process yields a variable number of drug-specific embeddings per patient, reflecting heterogeneity in treatment exposure. To synthesize these embeddings into a single patient-level representation, we computed the elementwise mean across all drug-specific embeddings.

Mean pooling was deliberately selected to summarize cumulative pharmacogenomic context in an order-invariant and parameter-free manner, thereby minimizing additional modeling assumptions and reducing the risk of overfitting given the modest cohort size and heterogeneous treatment histories. By avoiding attention-based or sequential aggregation mechanisms, this approach limits sensitivity to temporal noise and preserves a stable summary of interaction-level context suitable for downstream modeling.

We intentionally did not model treatment order, timing, or line of therapy at this stage. Incorporating such temporal dependencies would introduce additional trainable parameters and structural assumptions that are difficult to justify in small, real-world cohorts characterized by irregular and incomplete treatment trajectories. Accordingly, the objective of Stage 2 is to construct a stable representation of cumulative pharmacogenomic interaction context, rather than a full longitudinal treatment trajectory model. The resulting output is a fixed-dimensional vector compatible with standard downstream survival prediction architectures.

#### 2.3.3 Methodological Safeguards

Following Stage 1 training, all encoder parameters were frozen, and no further fine-tuning was performed using clinical data or survival outcomes. During Stage 2 inference, aggregation was fully deterministic and involved no trainable parameters. Consequently, the patient-level PGx embedding for any given individual was derived exclusively from observed genomic and chemical inputs available at the landmark time.

We confirmed that every patient in the cohort possessed at least one somatic mutation identified through targeted panel testing, enabling embedding generation for the complete cohort without reliance on imputation or placeholder values. Given identical mutation lists, drug identities (SMILES strings), encoder checkpoint, and aggregation procedures, the resulting patient-level PGx embeddings are fully reproducible across independent runs.

Collectively, these design choices ensure strict separation between representation learning and outcome modeling, minimize the risk of information leakage, and support transparent and reproducible transfer of experimental pharmacogenomic knowledge to the clinical setting.

### 2.4 Clinical Cohort and Baseline Modeling Framework

#### 2.4.1 Study Population

We constructed a retrospective clinical cohort of adult patients diagnosed with lung cancer at Mayo Clinic. To ensure high-fidelity molecular profiling relevant for pharmacogenomic modeling, inclusion was restricted to patients who underwent comprehensive genomic profiling via the FrontierOne CDx panel as part of routine clinical care. Eligible patients were further required to have complete electronic health record (EHR) documentation covering laboratory measurements and medication exposures within a predefined landmark window.

To ensure temporal consistency and minimize immortal time bias, a landmark time was defined as 90 days following the initial lung cancer diagnosis. All baseline features were extracted using data available at or before this landmark, and patients were subsequently followed to assess survival outcomes. Inclusion criteria strictly required data completeness across all three modalities (laboratory values, genomic profiling, and medication records) without reliance on data imputation. This rigorous selection process yielded a final analytically curated cohort of 184 patients, representing a fully aligned multimodal intersection that enables high-resolution integration across genomic, therapeutic, and laboratory domains.

#### 2.4.2 Baseline Multimodal Architectures

Baseline demographic, clinical, and modality-specific characteristics of the study cohort are summarized in Table 1. The mean age at diagnosis was 65.1 years (SD 10.4), and 51.1% of patients were female. At the landmark time point, 71.7% of patients presented with metastatic disease, and 45.7% had at least one major comorbidity, including cardiovascular, pulmonary, renal, metabolic, or connective tissue disorders.

**TABLE 1.** Demographics and Clinical Characteristics of the Study Cohort.

| Characteristic | Value at Landmark |
| --- | --- |
| **Demographics** |  |
| Age, years (Mean ± SD<sup>b</sup>) | 65.1 ± 10.4 |
| Sex, n (%) |  |
| Male | 90 (48.9) |
| Female | 94 (51.1) |
| **Clinical Characteristics** |  |
| Metastatic Disease, n (%) | 132 (71.7) |
| Key Comorbidities<sup>a</sup>, n (%) | 84 (45.7) |
| **Key Modality Summaries** |  |
| Laboratory Values |  |
| Albumin (g/dL), Mean ± SD | 3.8 ± 0.6 |
| Hemoglobin (g/dL), Mean ± SD | 11.7 ± 2.3 |
| Platelets (K/uL), Mean ± SD | 241.7 ± 119.0 |
| Genomic and Medication Profile |  |
| Mutated Genes per Patient, Median [IQR] | 5 [3-7] |
| Drug Classes per Patient, Median [IQR] | 9 [7-11] |
| **Outcome** |  |
| Died within 1-Year from Landmark, n (%) | 67 (36.4) |

<sup>a</sup> Key comorbidities include COPD, Coronary Artery Disease, Congestive Heart Failure, Chronic Kidney Disease, Diabetes, Cerebrovascular Disease, or Connective Tissue Disease diagnosed at or before the 90-day landmark. These clinical variables were used to characterize the cohort.
<sup>b</sup> SD: Standard Deviation; IQR: Interquartile Range. All data presented as n (%), unless otherwise specified

Genomic features were derived from the FrontierOne CDx panel, characterizing clinically actionable somatic alterations relevant to lung cancer. Patients harbored a median of five mutated genes (interquartile range [IQR], 3–7).

Medication exposure was characterized at the drug class level to capture broader therapeutic context rather than individual prescriptions. Patients were exposed to a median of nine distinct drug classes (IQR, 7–11) prior to the landmark time. The most prevalent drug classes reflected standard oncologic and supportive care regimens, with detailed distributions summarized in Table 1.

Laboratory features captured routinely measured clinical parameters commonly used in oncologic assessment. Table 1 reports summary statistics for key laboratory values, including albumin, hemoglobin, and platelet count (mean ± SD).

#### 2.4.3 Baseline Multimodal Architectures

To evaluate the contribution of pharmacogenomic representations across diverse modeling paradigms, we integrated PGx embeddings with four baseline architectures reflecting common approaches to multimodal EHR modeling. These architectures differ in how clinical information is represented and aggregated, enabling a systematic assessment of PGx complementarity across modeling assumptions.

- ENF (Expert-Engineered Numerical Features): A traditional tabular modeling approach utilizing curated clinical variables. Input features included normalized trajectories of 10 key laboratory tests, binary indicators for 27 medication classes, and binary indicators for somatic alterations across 271 genes. Missing values were encoded as zero to preserve sparsity patterns.
- CTE (Contextual Text Embedding): A semantic representation approach designed to incorporate biomedical context. Patient data were transformed into descriptive text profiles enriched with information from external knowledge bases, including DrugBank (22), Gene Ontology (23, 24), and KEGG (25). These profiles were encoded into fixed-length vectors using a pre-trained embedding model.
- E2E (End-to-End Transformer): A deep learning baseline using a long-context transformer architecture. ModernBERT (26) was fine-tuned directly on the same long-form textual profiles used by the CTE model, allowing the model to learn prognostic representations without explicit intermediate curation.
- GKC (Goal-oriented Knowledge Curators): A modular framework introducing an intermediate abstraction step. A large language model generated modality-specific, goal-oriented summaries prior to embedding, which were then concatenated to preserve complementary information across data sources.

The detailed explanations for baseline models are provided in prior stydy .

#### 2.4.3 Experimental Controls and Prediction Target

For all modeling architectures, pharmacogenomic embeddings were appended to baseline representations in a modular fashion to evaluate their complementary contribution under consistent experimental conditions. To prevent shortcut learning and ensure reliance on finer-grained biological and therapeutic signals, explicit tumor stage and metastatic status variables were excluded from all model inputs.

The primary prediction endpoint was one-year all-cause mortality following the landmark time. This time horizon aligns with the clinical context of the cohort, which is enriched for advanced-stage disease where short-term prognostication is clinically relevant. Model performance was assessed using repeated 5-fold cross-validation under identical data splits to ensure statistical rigor.

### 2.5 Experimental Setup: Testing the PGx Complementarity Representation Hypothesis

This section describes the experimental design used to evaluate the primary working hypothesis of this study: that externally learned pharmacogenomic embeddings act as a PGx Complementarity Representation whose contribution depends on the informational richness of the baseline EHR modalities. Specifically, we designed experiments to assess whether PGx embeddings provide larger relative benefit when baseline biological context is limited and reduced incremental benefit when such context is already present. To operationalize this hypothesis in a reproducible manner, we implemented two complementary experimental settings.

#### 2.5.1 Integration with Baseline Models

To assess whether the PGx embedding provides a robust and architecture-agnostic contribution, we evaluated its integration across four baseline survival modeling architectures (ENF, CTE, E2E, and GKC), each representing a distinct inductive bias for EHR representation. In all cases, the PGx embedding was concatenated with the baseline feature vector using a late-fusion strategy, preserving the original model structure and training procedure.

Each baseline model was trained and evaluated both with and without the PGx embedding under identical cross-validation folds and hyperparameter settings. This ceteris paribus design isolates the marginal contribution of pharmacogenomic information while controlling for architectural and optimization-related confounders. The objective of this experiment was to determine whether the PGx embedding yields consistent performance improvements across heterogeneous modeling strategies, as reported in Results Section 3.1.

#### 2.5.2 Modality Ablation: Validating the Gap-Filling Mechanism

To further characterize the modality-dependent behavior of the PGx embedding, we conducted a systematic ablation analysis across seven predefined combinations of EHR modalities, ranging from single-modality inputs (e.g., laboratory-only) to fully multimodal configurations. This analysis was designed to evaluate how the contribution of PGx embeddings varies as a function of baseline informational richness.

We formulated an a priori hypothesis that, if PGx embeddings encode mechanistic information that is partially orthogonal to standard EHR modalities, their relative contribution would be larger in modality-sparse settings and smaller in modality-rich settings. To quantify this effect, we defined the Performance Gain Ratio, which normalizes the absolute AUROC improvement observed for a given modality subset (**ΔAUC**_**subset**_) against the improvement observed in the full multimodal model (**ΔAUC**_**full**_):

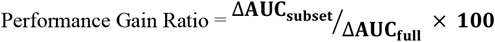

This normalized metric facilitates comparison across modality subsets with differing baseline performance levels and class distributions. AUROC was selected as the normalization metric because it provides a threshold-independent measure of discrimination that remains stable across heterogeneous feature configurations. Results from this analysis are presented in Results Section 3.2.

#### 2.5.3 Primary Performance Metrics and Statistical Testing

Model performance was primarily evaluated using the Area Under the Receiver Operating Characteristic curve (AUROC). These metrics were selected to provide threshold-independent assessments of discrimination that are well suited to one-year mortality prediction in high-risk cohorts. All experiments were conducted under a stratified five-fold cross-validation scheme.

To assess the statistical significance of performance differences between PGx-enhanced models and their corresponding baselines, we applied bootstrap resampling with 1,000 iterations. Performance improvements were considered statistically significant at a two-sided threshold of p < 0.05.

### 2.6 Patient-Level Feature Attribution and Robustness Analyses

#### 2.6.1 Patient-Level Aggregation of SHAP Evidence

To quantify the relative contribution of pharmacogenomic embeddings within multimodal models, we performed SHAP (SHapley Additive exPlanations) analysis (27). To measure modality-level contributions at the patient level, we utilized SHAP values of the top K features identified for each individual prediction. By restricting the analysis to the top K features, we standardized the evaluation scope to decision-critical evidence, enabling fair comparison despite substantial differences in feature dimensionality across modalities. For each patient, the absolute SHAP mass of the selected top K features was grouped by modality (Laboratory, Gene, Drug, and Pharmacogenomic [PGx]) and summarized as modality-specific contribution ratios, defined as the proportion of total absolute SHAP mass attributable to each modality within the top K set.

To mitigate variability arising from stochasticity in model training or data splitting, patient-level contribution ratios were averaged across 10 repeated 5-fold cross-validation runs. This procedure yielded a single stable contribution profile per patient, ensuring that each patient contributed one independent observation to subsequent statistical analyses. Patient-level comparisons between modalities were performed using Paired Wilcoxon signed-rank tests due to the non-Gaussian distribution of contribution ratios. This framework served as the basis for the primary patient-level comparisons presented in the main manuscript (Figure 4), with two-sided statistical significance assessed at p < 0.05.

#### 2.6.2 Robustness Assessment

To assess whether modality dominance patterns were robust to parameter choices and experimental variability, we conducted two complementary robustness analyses.

First, a K-sensitivity analysis was performed by varying the number of top-ranked features (K) across a predefined range (K = 10–50). For each K, patient-level contribution ratios were recomputed using the same aggregation and repeat-averaging procedure, allowing evaluation of the stability of modality contribution patterns across different scopes of decision evidence.

Second, a repeat-to-repeat stability analysis was conducted to characterize variability across independent cross-validation runs.

For each of the 10 repeats, modality-level contribution ratios were averaged across patients, yielding repeat-level mean contribution profiles for each modality. This analysis was used to assess the consistency of modality-level contribution patterns across runs, providing robustness characterization rather than primary inferential evidence.

#### 2.6.3 Mechanistic Decomposition and Confirmatory Analyses

Because contribution ratios are compositional and constrained to sum to one, a compositional log-ratio confirmatory analysis was performed to verify that observed modality differences were not artifacts of relative scaling. For each patient, log-ratios between PGx and each comparator modality were computed using repeat-averaged contribution profiles, and statistical significance was assessed using paired Wilcoxon signed-rank tests with Holm correction.

To further characterize the source of PGx dominance, a Shapley-based decomposition was applied to the difference in total top K contribution between PGx and Drug modalities. This framework partitioned the observed contribution gap into a count-driven component, reflecting differences in the selection frequency of modality-specific features among the top K set, and a mean-driven component, reflecting differences in the average SHAP magnitude of selected features. Patient-level bootstrap resampling was used to estimate confidence intervals for each component, enabling differentiation between dominance driven by the widespread accumulation of consistent signals versus dominance driven by a small number of extreme features.

### 2.7 Patient-Level Feature Attribution and Robustness Analyses

To assess whether the patient-level PGx Complementarity Representation encodes interpretable representational organization beyond aggregate predictive performance, we conducted a series of unsupervised analyses of the learned embedding space. The objective of this section is not to discover novel disease subtypes or to infer causal biological mechanisms, but rather to descriptively characterize patterns within the representation space and to evaluate whether these patterns align with known biological pathways and therapeutic exposures.

All analyses in this section were performed without access to clinical outcomes, survival times, disease stage, or histologic labels. The PGx representations analyzed here were generated exclusively from somatic mutation profiles and medication exposures available at the predefined landmark time, ensuring strict separation between representation analysis and outcome modeling.

#### 2.7.1 Unsupervised Clustering and Visualization

Unsupervised clustering was performed on the high-dimensional patient-level PGx embeddings using k-means clustering. The number of clusters (k = 3) was selected empirically to balance interpretability and descriptive clarity, rather than to optimize separation metrics or to infer an underlying ground-truth taxonomy. Cluster assignments were used solely for descriptive analyses and were not incorporated into any predictive models.

For visualization purposes, the high-dimensional embedding space was projected into two dimensions using t-distributed stochastic neighbor embedding (t-SNE). Dimensionality reduction was applied exclusively to facilitate qualitative inspection of representational geometry and did not influence clustering assignments or downstream analyses. Accordingly, any apparent separation observed in the projected space should not be interpreted as evidence of discrete clinical phenotypes, prognostic subgroups, or outcome-related stratification.

#### 2.7.2 Enrichment Analysis of Biological Pathways and Therapeutic Exposures

To characterize cluster-level associations, we performed enrichment analyses using two complementary feature sets: (i) administered anticancer therapies and (ii) Hallmark gene sets derived from patient genomic profiles. For each cluster, enrichment was assessed using Fisher’s exact test with false discovery rate correction to account for multiple testing.

In addition to statistical significance testing, we computed Cohen’s h as a directional effect size metric to summarize the relative magnitude and direction of over- or under-representation within each cluster. Cohen’s h was employed to facilitate symmetric interpretation of enrichment and relative depletion patterns across clusters, rather than to claim biological effect sizes or mechanistic relationships.

All enrichment results are interpreted as associative and descriptive, reflecting relative patterns within the learned representation space rather than causal links between molecular pathways, therapeutic exposures, and clinical outcomes.

#### 2.7.3 Interpretation Scope and Limitations

We emphasize that the unsupervised analyses presented here are exploratory in nature and are intended to support interpretation of the PGx Complementarity Representation. They are not designed to establish novel biological mechanisms, define clinical phenotypes, or propose alternative disease taxonomies. Observed patterns describe relative differences in representation-level associations and should be interpreted in light of the inherent limitations of unsupervised learning applied to heterogeneous real-world clinical data.

## 3. Result

To evaluate the potential utility of the PGx embedding as a PGx Complementarity Representation we investigated its capacity to mitigate the mechanistic gaps inherent in multimodal EHR data within a high-risk clinical setting. Given that our study cohort is heavily enriched for advanced and metastatic disease (72.7%), we focused on one-year mortality as the primary endpoint, representing a clinically critical horizon for risk stratification in real-world practice.

Across a series of controlled experiments, we systematically assessed whether the integration of pharmacogenomic knowledge could: (1) enhance survival prediction consistent across diverse modeling architectures, (2) provide complementary performance gains under modality-specific constraints, (3) serve as a salient driver of predictive signal relative to standard EHR features, and (4) identify latent patient subgroups reflecting biological coherence.

### 3.1 PGx embedding consistently improves survival prediction across all baseline architectures

To determine whether pharmacogenomic embeddings provide a robust prognostic signal beyond standard EHR features, we benchmarked one-year mortality prediction across four established survival architectures with distinct inductive biases (Fig. 2).

**Figure 1.**
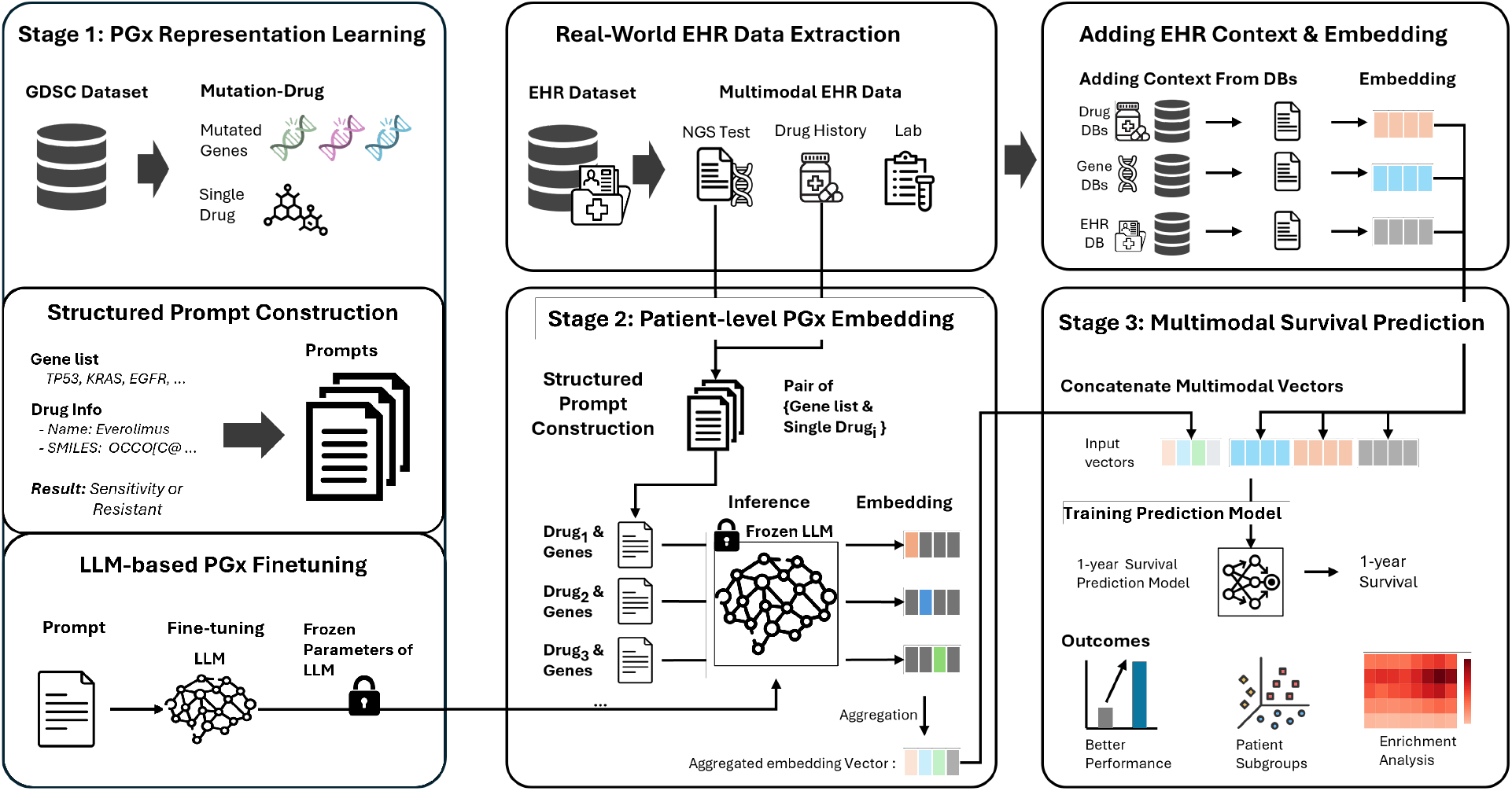
Overview of the PGx Complementarity Representation framework for transferring external pharmacogenomic knowledge into clinical survival prediction. The framework consists of three sequential stages. Stage 1 learns a mechanistic pharmacogenomic embedding from external experimental data (GDSC), where an LLM-based encoder is fine-tuned to model drug–mutation sensitivity relationships and subsequently frozen. Stage 2 transfers this representation to real-world patients by inferring drug–mutation embeddings from each patient’s genomic profile and treatment history, followed by mean pooling to construct a fixed-length, patient-level pharmacogenomic representation that enables zero-shot encoding without outcome leakage. Stage 3 integrates the resulting PGx embedding with multimodal EHR features (laboratory values, genomics, and medications) for downstream survival prediction and interpretation. This design enables architecture-agnostic performance gains while preserving biological coherence and interpretability, operationalizing pharmacogenomic knowledge as a complementary signal rather than a replacement for standard clinical data.

**Figure 2.**
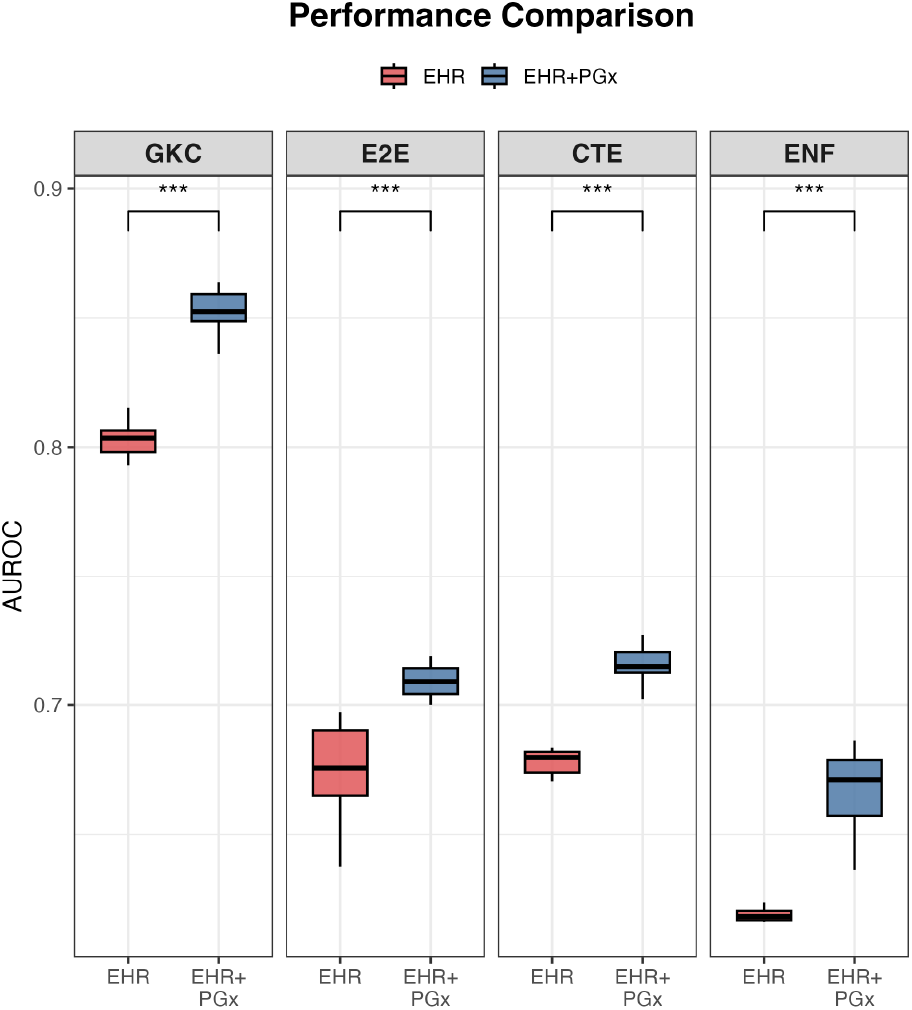
Pharmacogenomic embeddings consistently improve survival prediction across diverse modeling architectures. Comparison of AUROC for one-year mortality prediction across four baseline architectures (ENF, CTE, E2E, and GKC) using standard EHR features alone (red) versus EHR augmented with pharmacogenomic (PGx) embeddings (blue). Across all architectures, incorporation of PGx embeddings results in statistically significant performance gains (***p < 0.001), indicating robust and architecture-agnostic improvement.

The integration of PGx embeddings was associated with statistically significant performance gains across every evaluated baseline architecture (p < 0.05 for all comparisons), regardless of model complexity. Rather than offering negligible additive value, the inclusion of pharmacogenomic representations consistently extended predictive performance beyond the baseline levels achieved with standard multimodal EHR features alone.

Notably, these improvements were observed not only in simpler models but also in our strongest baseline, the GKC framework, where the mean AUROC increased from 0.803 to 0.852 (95% CI: 0.847–0.857). This universality suggests that the PGx embedding captures complementary biological signals that are not explicitly resolved by traditional clinical variables. The stability of these gains across folds and metrics supports the utility of the PGx embedding as a consistent source of prognostic information.

Given the high prevalence of metastatic disease in this cohort, we further investigated whether these improvements might be driven by coarse clinical proxies. We conducted stratified validation analyses to compare the model against a metastasis-based naïve rule. The PGx-enhanced model demonstrated superior discrimination compared to the naïve rule and successfully stratified one-year mortality risk even within the metastatic-only subgroup (p < 10^−6^). Collectively, these analyses indicate that the observed performance gains likely reflect finer-grained biological and therapeutic signals rather than an implicit reliance on disease stage.

### 3.2 PGx-driven improvements persist across multiple modality combinations in systematic ablation analyses

To investigate how pharmacogenomic (PGx) embeddings contribute across different modality settings, we conducted a systematic modality ablation analysis examining how PGx information interacts with diverse combinations of EHR-derived features (Fig. 3). In this analysis, complementarity is operationally defined at the representation level, such that the marginal contribution of PGx embeddings varies according to the informational richness of the baseline EHR modalities. Within this framework, PGx embeddings are treated as a derived, interaction-aware representation appended to existing features, rather than as a replacement for observed genomic or therapeutic variables.

**Figure 3.**
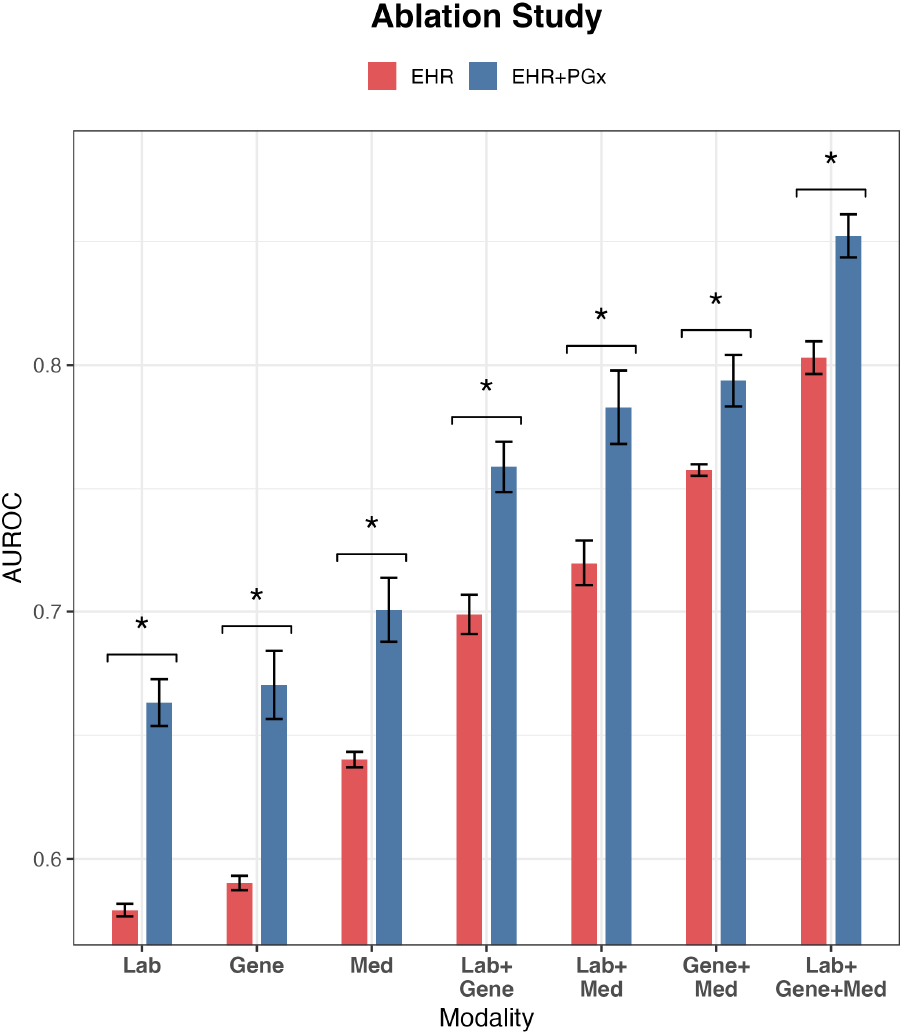
PGx embeddings yield consistent and context-dependent gains across modality ablations. Comparison of AUROC improvements across seven EHR modality combinations. Integration of PGx embeddings significantly improved AUROC in all settings (p < 0.05). Relative gains were largest in modality-sparse configurations (e.g., Lab-only) and attenuated in biologically enriched models (e.g., Gene+Drug), consistent with the interpretation that the PGx Complementarity Representation provides context-dependent, non-redundant representational augmentation, rather than replacing existing clinical features.

Across all seven evaluated modality combinations, the inclusion of PGx embeddings was associated with statistically significant improvements in AUROC (p < 0.05 for all configurations). Importantly, the magnitude of these improvements exhibited a clear dependency on baseline modality composition. In modality-sparse settings, PGx embeddings yielded the largest relative gains. Specifically, the Lab-only model demonstrated the greatest relative improvement when normalized to the full multimodal reference gain (ΔAUROC = +0.084, corresponding to 171% of the reference improvement). This pattern suggests that the PGx Complementarity Representation provides additional representational context when baseline EHR modalities encode limited biological information.

In contrast, in biologically enriched configurations that explicitly incorporate genomic and therapeutic information, the relative gains associated with PGx embeddings were attenuated. For example, the Gene+Drug model exhibited a smaller relative improvement (73%; ΔAUROC = +0.036), consistent with partial representational overlap between explicitly encoded genomic features and the PGx Complementarity Representation. Notably, performance gains remained statistically significant even in these enriched settings, indicating that the PGx representation contributes non-redundant information beyond that captured by standard clinical features alone.

We emphasize that this ablation analysis reflects a representation-space augmentation setting, rather than an information-budget– controlled comparison. Accordingly, these results are not intended to suggest that PGx embeddings replace observed genomic or therapeutic features. Instead, they indicate that the marginal impact of the PGx Complementarity Representation is context-dependent, with the strongest relative contribution observed when baseline EHR modalities encode limited biological context.

To further address attribution concerns, we compared PGx-enhanced models against a baseline using raw gene and drug identifiers without external biological context (ENF). The PGx representation consistently outperformed this baseline (AUROC 0.652 vs. 0.601), supporting the effective transfer of interaction-aware representational signals learned from external pharmacogenomic data. Furthermore, even when explicit Gene and Drug features were included in the baseline, the addition of PGx embeddings yielded additional performance improvements (AUROC 0.794), indicating that observed gains cannot be attributed to simple information reuse.

Collectively, these results support the interpretation that the PGx Complementarity Representation functions as a context-dependent, non-redundant augmentation within multimodal EHR models. Its relative contribution is amplified in modality-sparse configurations and attenuated—but not eliminated—in biologically rich settings, consistent with a representation-level notion of complementarity rather than explicit mechanistic replacement.

### 3.3 PGx embedding emerges as a primary source of predictive signal

To quantify modality-level contributions at the patient level, we summarized each patient’s top-K SHAP evidence by computing the contribution ratio of each modality (Lab, Gene, Drug, PGx) within the selected top-K features (Fig. 4). To obtain stable patient-level estimates, these ratios were averaged across 10 repeated 5-fold cross-validation runs. Using K = 10 as the primary setting to capture decision-critical evidence, PGx accounted for the largest share of contributions (median 0.413, IQR [0.327, 0.528]) compared with Drug (median 0.219), Gene (median 0.192), and Lab (median 0.144), with all contrasts remaining statistically significant (p < 0.001 for all). Patient-level paired comparisons further confirmed that PGx contribution ratios were consistently higher than each comparator modality (median differences ranging from 0.181 to 0.217). Sensitivity analyses demonstrated that this dominance was robust to hyperparameter selection, as the PGx contribution ratio increased as K expanded from 10 to 50, and the run-level difference between PGx and Drug remained consistently positive across all repeated runs (mean difference 0.182, SD 0.041).

To further characterize the structure underlying this attribution pattern, we performed compositional and Shapley-based confirmatory analyses. Log-ratio analysis confirmed that PGx contributions remained significantly higher than all comparator modalities after accounting for compositional constraints (all p < 0.001). We then decomposed the PGx contribution advantage into selection-frequency (count-driven) and effect-magnitude components. This analysis showed that the PGx advantage was primarily driven by the frequent inclusion of PGx features among top-ranked decision evidence rather than by rare, extreme-magnitude effects. This count-driven dominance corresponds to a pattern in which attribution for standard clinical features is distributed across correlated variables, whereas PGx features are repeatedly selected as distinct interaction-level signals.

### 3.4 Unsupervised Characterization of Latent Bio-Clinical Phenotypes

Taken together, these exploratory analyses suggest that the PGx embedding space captures biologically and therapeutically coherent structure defined by both enrichment and depletion patterns. While qualitative in nature, these findings provide supportive evidence that the proposed PGx Complementarity Representation reflects underlying mechanistic relationships linking pharmacogenomic context and therapeutic exposure

To examine whether the PGx Complementarity Representation encodes interpretable organization beyond aggregate predictive performance, we conducted a series of unsupervised analyses on the learned patient-level embedding space. These analyses were exploratory and descriptive in nature, intended to support interpretation of the representation rather than to define novel clinical phenotypes or to infer causal biological mechanisms.

Patient-level PGx embeddings were analyzed without access to survival outcomes, disease stage, or histologic labels. Unsupervised clustering was performed using k-means on the high-dimensional embedding vectors, with the number of clusters (k = 3) selected empirically to facilitate interpretability and descriptive clarity rather than to optimize separation metrics or to infer an underlying ground-truth taxonomy. To enable qualitative inspection of embedding geometry, representations were projected into two dimensions using t-distributed stochastic neighbor embedding (t-SNE). Dimensionality reduction was used solely for visualization purposes and did not influence clustering assignments or downstream analyses.

Visualization of the projected embedding space revealed apparent organization into three latent groups (Fig. 5A). Because t-SNE is a visualization technique rather than a statistical test, observed separation in the projected space should not be interpreted as evidence of discrete clinical subtypes. Instead, these patterns motivated subsequent descriptive analyses of clinical and biological associations characterizing each cluster.

**Figure 4.**
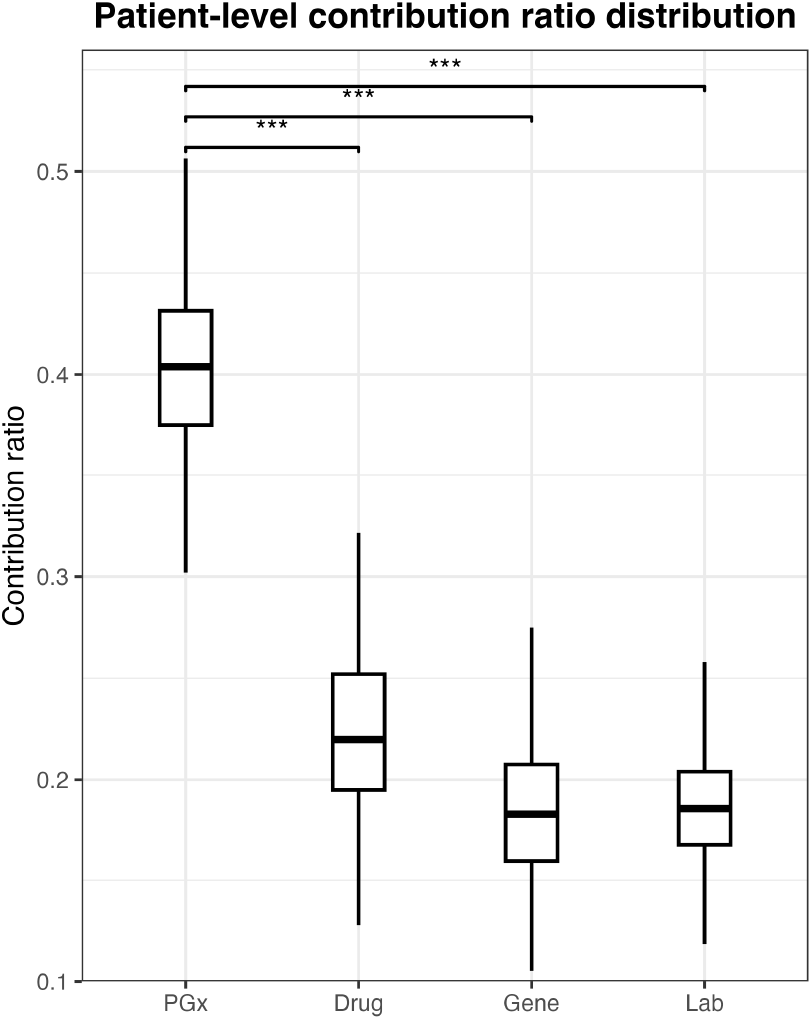
Patient-level distribution of modality contribution ratios. Box plots summarize patient-level contribution ratios for each modality (Lab, Gene, Drug, PGx) computed from top K SHAP evidence (K = 10). For each patient, contribution ratios were averaged across 10 repeated 5-fold cross-validation runs to obtain a stable estimate. Boxes indicate the median and interquartile range (IQR). PGx exhibits the largest contribution ratio (median 0.413, IQR [0.327, 0.528]) compared with Drug (median 0.219), Gene (median 0.192), and Lab (median 0.144). Brackets denote paired Wilcoxon tests comparing PGx with each comparator modality (p < 0.001 for all).

**Figure 5.**
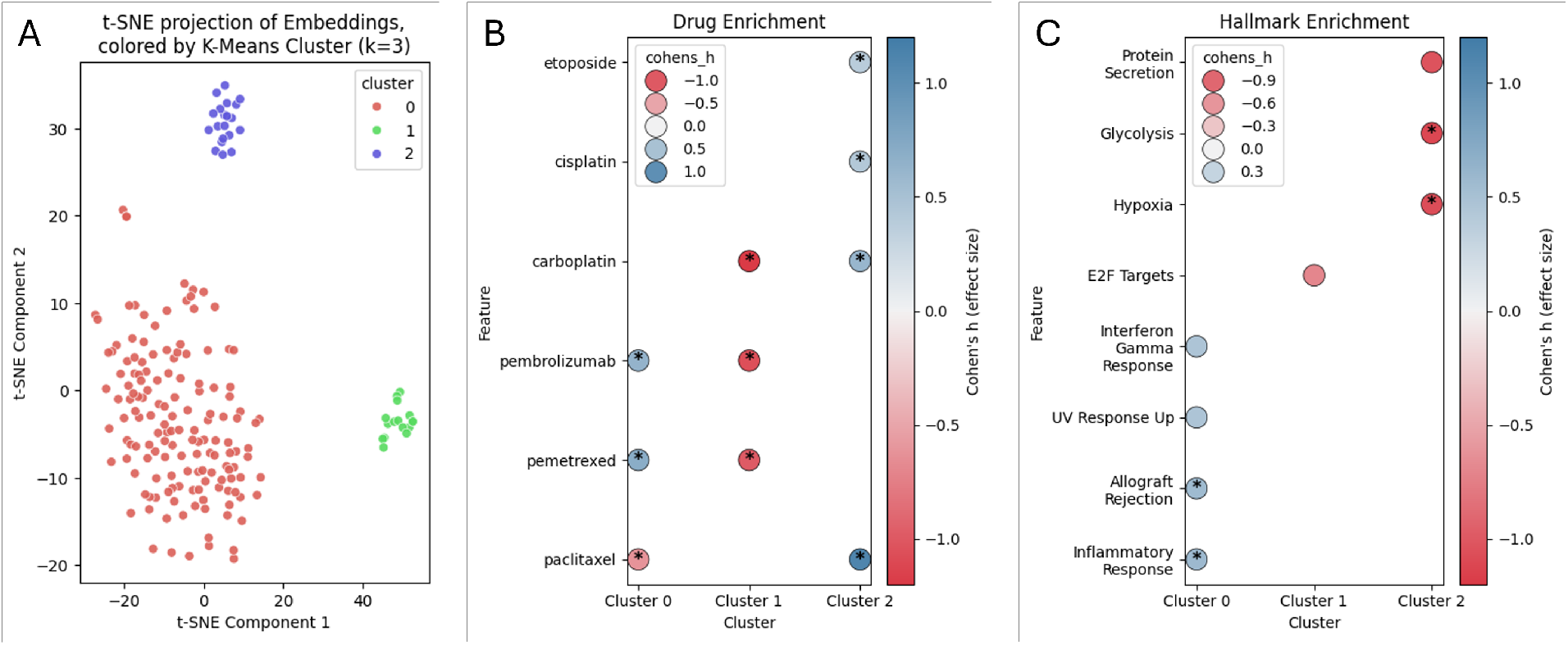
Unsupervised analysis of PGx embeddings visualizes biologically and therapeutically coherent patient subgroups. (A) t-SNE projection of patient-level PGx embeddings colored by K-means clustering (k = 3). The embedding space shows spontaneous separation into three distinct latent subgroups without the use of clinical outcome labels or histologic annotations. (B) Drug enrichment analysis illustrating cluster-specific treatment patterns. Directional effect sizes are quantified using Cohen’s h, where positive values indicate enrichment and negative values indicate relative depletion compared to the remaining cohort. Circles denote drugs with nominal significance (p < 0.05), and starred circles indicate associations surviving multiple testing correction (FDR q < 0.15). (C) Hallmark pathway enrichment analysis highlighting coherent biological programs. Cluster 0 shows positive enrichment for immune-related pathways, whereas Cluster 1 is characterized by the systematic depletion of proliferative signals consistent with a quiescent phenotype. Collectively, these results suggest that PGx embeddings capture underlying bio-clinical structure by aligning molecular pathway activity with therapeutic exposure in a directionally coherent manner.

To characterize cluster-level associations, we conducted enrichment analyses using two complementary feature sets: administered anticancer therapies and Hallmark gene sets derived from patient genomic profiles. Associations were evaluated using Fisher’s exact test with false discovery rate adjustment. To facilitate symmetric interpretation of both enrichment and relative depletion patterns, we additionally computed Cohen’s h as a directional effect size metric. In this context, Cohen’s h was used to summarize the direction and magnitude of relative over-or under-representation within clusters, rather than to claim biological effect sizes or mechanistic relationships.

Distinct and consistently patterned associations were observed across clusters (Fig. 5B–C). One cluster exhibited relative enrichment for immune-related Hallmark pathways, including interferon-gamma signaling, alongside overrepresentation of immune checkpoint inhibitors such as pembrolizumab. This pattern is consistent with alignment between immune-related molecular features and corresponding treatment exposures. A second cluster showed relative enrichment for cytotoxic chemotherapy agents, including paclitaxel and carboplatin, with less pronounced pathway-level associations, suggesting a treatment-dominant representational pattern rather than convergence on a single biological axis.

In contrast, the remaining cluster was characterized by a relative depletion pattern, with negative Cohen’s h values observed across both therapeutic exposures and proliferative Hallmark pathways. Rather than indicating the absence of meaningful organization, this pattern reflects a consistent representational profile marked by lower relative association with intensive treatment regimens and proliferative signaling programs. Importantly, these observations describe relative differences within the embedding space and should not be interpreted as clinical risk stratification, prognostic subgrouping, or outcome-related categorization.

Collectively, these exploratory analyses indicate that the PGx Complementarity Representation captures structured bio-clinical association patterns that align with known classes of treatment exposure and pathway activity. While these findings do not establish causal mechanisms or novel disease subtypes, they provide supportive interpretability evidence that the learned representations encode meaningful biological and therapeutic context beyond aggregate predictive performance.

## 4. Discussion

In this study, we investigated whether externally learned pharmacogenomic representations can address a persistent limitation of multimodal EHR based survival modeling, namely the absence of explicit mechanistic links between genomic mutations and therapeutic response (28). By conceptualizing the PGx embedding as a PGx Complementarity Representation, we evaluated a two-stage framework that transfers drug–gene interaction patterns learned from large-scale experimental screens into real-world lung cancer survival prediction. Across a series of controlled analyses, our findings indicate that this representation provides a consistent and stable prognostic signal beyond standard EHR features.

The present study is situated within a broader institutional effort to develop high-fidelity genomic–EHR integration for clinical outcome modeling. Prior work established a pan-cancer genomic– EHR linkage infrastructure with standardized patient matching and harmonized clinical representations, providing a reliable parent data environment for disease-specific analyses [AMIA Joint Summits on Translational Science, 2020]. Building on this foundation, we deliberately transitioned to a lung cancer–specific cohort defined by comprehensive genomic profiling and strict temporal alignment of laboratory and medication data at a fixed landmark.

Importantly, the cohort analyzed here reflects a prespecified multimodal intersection designed to support unbiased representation learning across heterogeneous clinical modalities. While this curated lung cancer cohort has previously served as a stable experimental setting for baseline predictive modeling under an identical task definition, the present work extends beyond performance evaluation by systematically examining modality complementarity and the biological structure encoded by pharmacogenomic representations.

This design choice constitutes a key strength of the study. By prioritizing multimodal fidelity and temporal consistency over cohort size, we were able to isolate the scientific effect of externally learned pharmacogenomic embeddings on survival prediction and interpret their contribution relative to existing EHR modalities. As a result, the observed performance gains and representation-level insights can be attributed to modality-specific information transfer rather than artifacts of missing data, temporal misalignment, or post hoc sample selection. These findings provide a principled framework for understanding when pharmacogenomic representations are most informative within multimodal clinical models.

A key observation of this work is that the contribution of the PGx embedding is not uniform across modeling contexts. Instead, its impact depends on the informational richness of the baseline modalities. Through systematic modality ablation, we observed that relative performance gains were most pronounced in settings where explicit biological context was limited, such as laboratory-only models, while gains diminished in configurations already containing genomic and medication information. This pattern is consistent with the interpretation that the PGx embedding captures information that is partially overlapping with, but not redundant to, existing EHR modalities (29). Rather than acting as a generic feature augmentation, the embedding appears to provide targeted biological context where such information is otherwise sparse.

The present findings characterize complementary aspects of how PGx embeddings operate within a multimodal predictive framework. Ablation analyses demonstrate that the incremental value of PGx depends on the informational richness of the baseline feature set, with the strongest augmentation effects observed in data-sparse settings. These results delineate the conditions under which PGx contributes additional predictive signal relative to the available clinical context, highlighting its role as a context-dependent augmentation rather than a standalone substitute.

In contrast, attribution analyses examine how predictive evidence is allocated once all modalities are jointly available, focusing on marginal contribution to patient-level decision-making rather than on isolated predictive adequacy. In this setting, feature attribution reflects the uniqueness of information rather than its completeness. Standard EHR modalities often encode overlapping aspects of disease burden, resulting in their predictive contribution being distributed across correlated features. By comparison, the PGx embedding captures interaction-level mechanistic signals that are not redundantly represented elsewhere, leading to concentrated and consistently selected attribution at the patient level. Together, these findings indicate that PGx embeddings function not as a replacement for explicit clinical variables, but as an orthogonal decision axis that refines individual risk stratification once the clinical baseline is established.

Beyond aggregate performance metrics, we further examined whether the learned representations exhibit internal structure consistent with known biological and therapeutic patterns. Unsupervised analyses revealed that patient embeddings organized into distinct subgroups associated with characteristic pathway enrichments and treatment exposures. Notably, these patterns emerged without the use of explicit histologic or molecular subtype labels. While such clustering analyses are inherently exploratory, the observed alignment between pathway activity and therapeutic regimens suggests that the PGx embedding retains biologically meaningful organization, supporting its interpretability beyond predictive accuracy alone (30).

Given the advanced disease composition of our cohort, we specifically focused on one-year mortality as a clinically relevant prediction horizon. In this setting, an important concern is whether predictive models rely disproportionately on coarse clinical proxies, such as metastatic status (31). To address this, we conducted stratified validation analyses demonstrating that PGx-enhanced models retain discriminative ability within both metastatic and non-metastatic strata and outperform a metastasis-based naïve rule. These results suggest that the observed performance gains are not solely attributable to implicit encoding of disease stage but instead reflect finer-grained biological and therapeutic information captured by the embedding.

Several limitations of this study should be acknowledged. First, the analysis was conducted using a single-center cohort with a deliberately restricted sample size, reflecting the requirement for comprehensive genomic profiling and complete multimodal alignment at a fixed landmark. While this design prioritizes internal validity and controlled assessment of representation learning, external validation in larger and more diverse populations will be necessary to evaluate generalizability across broader clinical settings. Second, the pharmacogenomic representations were derived from in-vitro cell-line data, which may not fully recapitulate in-vivo tumor behavior (32). In addition, drug sensitivity was represented using binary labels derived from GDSC-defined thresholds, which necessarily abstract continuous IC50 measurements and may not fully capture finer-grained dose–response variability across cell lines. Although our results suggest successful transfer of these signals to patient-level outcomes, future work incorporating patient-derived models or longitudinal real-world pharmacologic data could further strengthen this connection. Finally, this study was retrospective in nature; prospective evaluation will be required to determine whether PGx-informed risk stratification can meaningfully influence clinical decision-making in practice.

## 5. Conclusion

We present a reproducible informatics framework for transferring external pharmacogenomic knowledge into EHR-based survival modeling. By operationalizing pharmacogenomic information as a PGx Complementarity Representation, our approach enables interaction-aware augmentation of clinical models while remaining independent of outcome supervision and additional model complexity. Across systematic evaluations, this representation provides consistent, context-dependent improvements and supports interpretable association patterns aligned with known biological and therapeutic contexts. Together, these findings illustrate how externally learned biological knowledge can be integrated into real-world clinical prediction frameworks in a principled and scope-disciplined manner.

## Data Availability

The data used in this study were derived from electronic health records at Mayo Clinic and contain protected health information. Due to patient privacy considerations and institutional restrictions, these data are not publicly available. 
Aggregated results and methodological details sufficient to reproduce the analyses are provided in the manuscript.

## Notes

### Competing Interest Statement

The authors have declared no competing interest.

### Funding Statement

This work was supported by institutional research funding from Mayo Clinic. The funding sources had no role in the study design, data collection, analysis, interpretation of results, or manuscript preparation.

### Author Declarations

The Institutional Review Board of Mayo Clinic gave ethical approval for this work.

